# COVIDTrach; the outcomes of mechanically ventilated COVID-19 patients undergoing tracheostomy in the UK: Interim Report

**DOI:** 10.1101/2020.05.22.20104679

**Authors:** COVIDTrach collaborative, NJI Hamilton, T Jacob, AGM Schilder, A Arora, MM George, F Green, E Jackson, J Goulder, N Kumar, C Schilling, S Laha, I Ahmad, B McGrath, MA Birchall, NS Tolley, G Sandhu, T Tatla, N Sharma, P Stimpson, P Andrews, P Surda, A Takhar, C Xie, E Kamta Bhargava, C Tornari, M Verkerk, M Kelly, J Collins, D Pennell, N Amin, D Ranford, C Al-Yaghchi, L Ritchie, M Jaafar, R Mistry, M Rouhani, M Ashcroft, N Cereceda-Monteoliva, A Holroyd, J Ng, K Ghufoor, E Warner, H Drewery, J Hadley, R Bhandari, N Bhatti, H O’mahoney, S Shepherd, H Wilson, M Griffiths, A Rovira, T Munroe-Gray, D Dawson, P Sethukumar, I Ekpemi, RR Bance, K Karamali, N Glibbery, C Walker, K Valchanov, L Bates, S Saha, C Smart, T Magos, A Loizidou, M Lee, D Allin, R Kumar, B Cosway, R Glore, E Omakobia, G Tattersall, B Hill, W Udall, S Khwaja, R Anmolsingh, C Smyth, B Al-Dulaimy, K Kapoor, S Sirajuddin, S Fang, F Van-Damme, D Bondin, D Thorley, D Nair, S Kandiah, C Davies Husband, C Barrera-Groba, N Seymour, S Mahalingam, E Leakey, S Okhovat, H Buglass, E Tam, U Sheikh, S Suresh, J Westwood, J Smith, M Celinski, S Shahidi, K Jolly, M Osborne, J Fussey, P Kirkland, J Staufenberg, R Vasanthan, S Ladan, P Paul, P Tsirevelou, V Ratnam, M Anwar, A Pericleous, J Bates, R Moorthy, P Bothma, S Meghji, O Judd, T Ali, T Stubington, A Kumar, W Parker, T Davis, A Burgess, A Tsagkovits, S Winter, T Hunt, A Vijendren, V Venkatachalam, M Lechner, D Chandrasekharan, A Arya, R Brown, V Srinivasan, M Junaid, R Temple, R Pinto, U Nagalotimath, R Sheikh, C Cook, J Lunn, B Ranganathan, N Mani, H Saeed, S Linton, R Stewart, S Nakagawa, H Turner, J Whiteside, J Whiteside, F Cooper, J Collier, P Ward, C Lockie, L Lignos, A Courtney, T Browning, O Mulla, N Stobbs, A Alegria, S Starnes, A Thompson, J Whittaker, A Hassan, M Cameron, A Walker, L Leach, P Gill, L McCadden, S Baker, S Sanyal, S Wilkinson, R Siau, N Vallabh, Emma Riley, Ahmad K. Abou-Foul

## Abstract

COVIDTrach is a UK multidisciplinary collaborative project that aims to evaluate the outcomes of tracheostomy in COVID-19 patients. An invitation to participate in an online survey tool (REDCap) was disseminated to all UK NHS departments involved in tracheostomy in mechanically ventilated COVID-19 patients. Fifty-two percent (n=219/465) of patients who had undergone tracheostomy and were still alive, had been successfully weaned from mechanical ventilation at the point of completing the survey. The all cause in-hospital mortality following tracheostomy was 12% (n=62/530), with 3% of these (n=2/62) due to tracheostomy related complications and the remaining deaths due to COVID-19 related complications. Amongst 400 cases submitting data two weeks after the tracheostomy, no instance of COVID-19 infection amongst operators was recorded. This interim report highlights early outcomes following tracheostomy in mechanically ventilated COVID-19 patients. Future reporting from COVIDTrach will include more detailed analysis at later timepoints using comparator groups in order to provide a more comprehensive assessment of tracheostomy in COVID-19.

## Correspondence

COVIDTrach is a UK multidisciplinary collaborative project that evaluates the outcomes of tracheostomy in patients diagnosed with COVID-19 who are receiving invasive mechanical ventilation. In parallel, data is collected on the use of personal protective equipment (PPE) and rates of COVID-19 infection amongst operators. Between 6^th^ April and 11^th^ May 2020, data was received on 564 tracheostomies from 78 UK NHS hospitals. Results are given in brackets as a fraction of results received (n=results/number of results received).

The majority of patients were male (n=405/563, 72%) and BMI ranged from 18.5 to <25 (22%), 25 to <30 (35%), 30 to <40 (35%) and >40 (8%) (data available in 426 cases). The number of days from intubation (day 0) to tracheostomy ranged from 0-35 (median 16, IQR 13, 22) (data available in 543 cases). Prior to tracheostomy, the median Fi02 was 40% (IQR 30, 45) (data available in 555 cases) and the median PEEP was 8 (IQR 6, 10) (data available in 539 cases). An open method of tracheostomy was used in 58% of cases (n=323/560), a percutaneous method in 39% (n=217/560) and a hybrid method was used in 3% (n=20/560). A negative pressure environment was used in 10% of cases (n=55/530).

Fifty-two percent (n=219/465) of COVID-19 patients who had undergone tracheostomy and were still alive had been weaned from mechanical ventilation at the point of completing the survey (Table 1). At the point of survey, 38% (n=169/450) of patients who had undergone a tracheostomy had been discharged from intensive care. The all-cause in-hospital mortality following tracheostomy in COVID-19 patients was 12% (n=62/530) with two deaths attributed to post-operative tracheostomy complications and the other 60 (97%) recorded as “COVID-19 related”. As many of the patients are yet to complete their critical care, the mortality and weaning rates are likely to change with time. The success in tracheostomy decannulation and discharge from hospital will be evaluated in future reports.

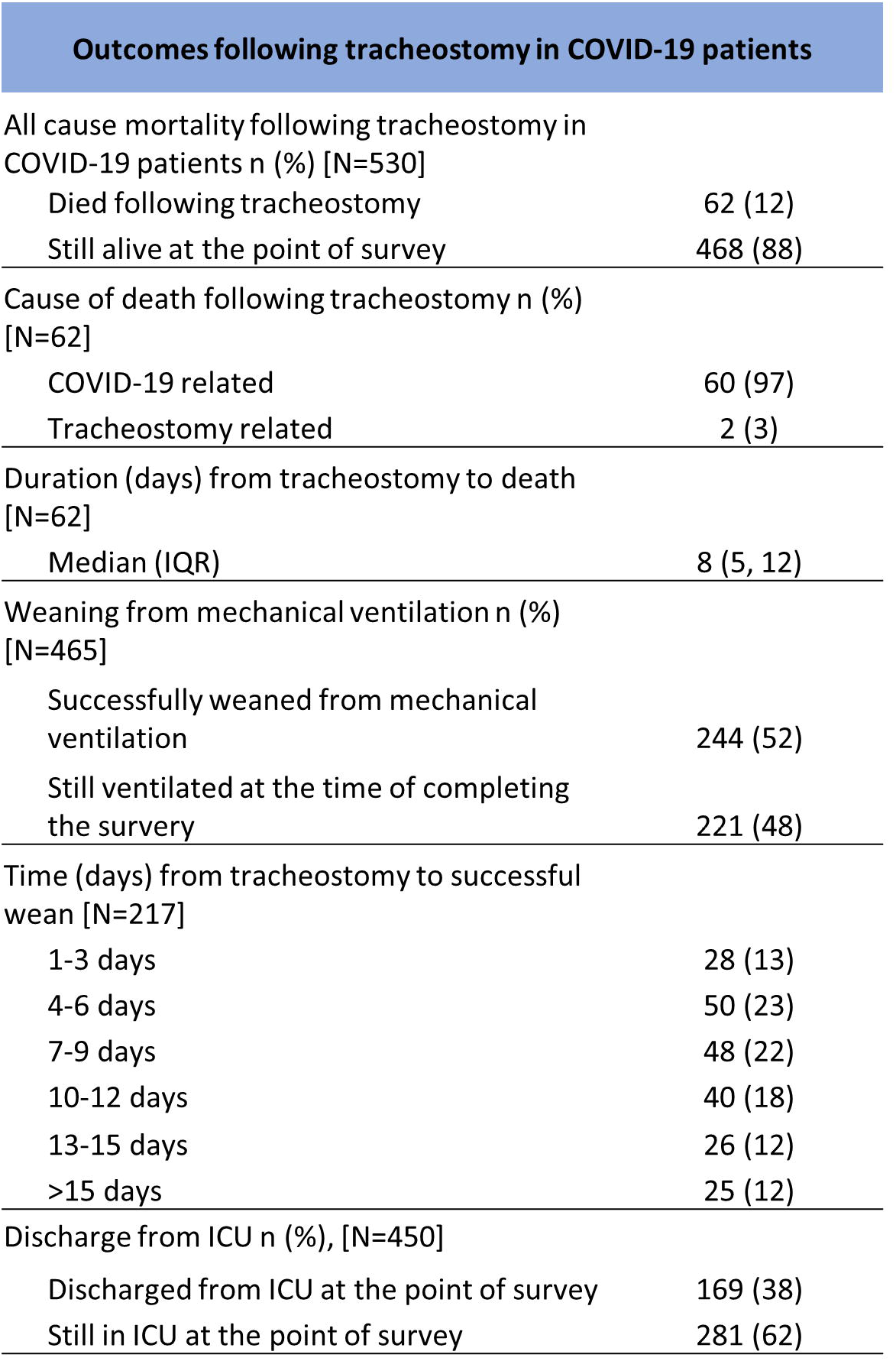

|  |  |
| --- | --- |
| Died following tracheostomy | 62 (12) |
| Still alive at the point of survey | 468 (88) |
| Cause of death following tracheostomy n (%) [N=62] |  |
| COVID-19 related | 60 (97) |
| Tracheostomy related | 2 (3) |
| Duration (days) from tracheostomy to death [N=62] |  |
| Median (IQR) | 8 (5, 12) |
| Weaning from mechanical ventilation n (%) [N=465] |  |
| Successfully weaned from mechanical ventilation | 244 (52) |
| Still ventilated at the time of completing the survey | 221 (48) |
| Time (days) from tracheostomy to successful wean [N=217] |  |
| 1-3 days | 28 (13) |
| 4-6 days | 50 (23) |
| 7-9 days | 48 (22) |
| 10-12 days | 40 (18) |
| 13-15 days | 26 (12) |
| >15 days | 25 (12) |
| Discharge from ICU n (%), [N=450] |  |
| Discharged from ICU at the point of survey | 169 (38) |
| Still in ICU at the point of survey | 281 (62) |

Adequate PPE should be viewed as mandatory for tracheostomy in COVID-19 patients due to the significant potential for aerosol generation.^1,2^ In all cases (n=545/545), operators used either an FFP3 mask or Powered Air Purifying Respirator (PAPR). Additional PPE used involved either a face visor or hood with face shield in 99% of cases (n=538/545), a disposable gown in 97% (n=527/545) and double gloves in 90% (n=490/545). The question “Did any of the operators test positive for COVID-19 within two weeks of the procedure”, was answered by 71% (n=400/564) and all confirmed that no operators had become COVID-19 test positive within two weeks of the procedure. Whilst this finding is reassuring, it is open to potential reporting bias and does not account for the remaining cases that are yet to reach the two-week time point.

The number of COVID-19 PCR tests performed prior to tracheostomy ranged from 1 to 12 (median 1, IQR 1,2). The COVID-19 test was positive in 86% (n=443/503) of patients prior to tracheostomy with the length of time from the last test to the day of surgery recorded as median 14 days (IQR 7,19). The role of identifying PCR test status in COVID-19 patients ahead of tracheostomy is unclear. ICU patients can remain test positive for several weeks after the onset of symptoms,^3,4^ and whether the detection of viral RNA by PCR predicts risk of infectivity to operators and other health care professionals is uncertain. Delaying tracheostomy to achieve negative tests is likely to prolong endotracheal ventilation and thus defer the potential benefits of tracheostomy whilst increasing the risk of complications relating to endotracheal intubation.

## Data Availability

All data is saved on a secure server at University College London.

## Data collection

Study data were collected and managed using REDCap electronic data capture tools hosted at University College London. REDCap (Research Electronic Data Capture) is a secure, web-based application designed to support data capture for research studies, providing: 1) an intuitive interface for validated data entry; 2) audit trails for tracking data manipulation and export procedures; 3) automated export procedures for seamless data downloads to common statistical packages; and 4) procedures for data integration and interoperability with external sources.

## Conflicts of interests

No conflicts to declare.

## Funding source

COVIDTrach is supported by the UCL / Wellcome Trust COVID-19 rapid response fund. Nick Hamilton is supported by a National Institute of Health & Research (NIHR) lectureship, the Academy of Medical Sciences Starter Grant and the Royal College of Surgeons. Anne GM Schilder is an NIHR Senior Investigator and Director of the NIHR UCLH BRC Hearing Theme; the research of her UCL Ear Institute evidENT team is supported by the National Institute for Health Research (BRC, ARC, CRN, PGfAR, RfPB), Wellcome Trust, RCSEng, and EU Horizon2020.

## Notes

### Competing Interest Statement

The authors have declared no competing interest.

### Author Declarations

NHS Health Research Authority

